# Overview on COVID-19 outbreak indicators across Brazilian federative units

**DOI:** 10.1101/2020.06.02.20120220

**Authors:** Elvira Maria Guerra-Shinohara, Simone Schneider Weber, Clóvis Paniz, Guilherme Wataru Gomes, Eduardo Jun Shinohara, Tiago Borges Ribeiro Gandra, Indiara Correia Pereira, Karine Gomes Jarcem, Renato Ferreira de Almeida Zanre, Acácia Gimenez Barreto, Alessandro Diogo De Carli

## Abstract

**Background:** The 2019 coronavirus disease pandemic (COVID-19) spread rapidly across Brazil. The country has 27 federative units, with wide regional differences related to climate, lifestyle habits, socioeconomic characteristics and population density. Therefore, we aimed to document and monitor the increase in COVID-19 cases across each federative unit in Brazil, by tracking its progression from inception to 15 May 2020.

**Methods:** Observational study.

**Results:** The first confirmed COVID-19 case in the country was notified in São Paulo on 26 February, while the first death occurred on 17 March, in Rio de Janeiro. Since then, there has been a dramatic increase in both confirmed cases and deaths from the disease. São Paulo, in the Southeast region, was initially considered the COVID-19 epidemic epicentre in Brazil. However, 10 states in the North and Northeast regions were ranked among the 14 highest incidences (over 100 cases per 100,000 people) observed on 15 May. Higher incidence rates (>100 cases per 100,000) were associated to higher rates of inadequate water supply and sewerage (OR, 5.83 (95% CI, 1.08–29.37, P=0.041)). North and Northeast states with the highest social vulnerability index scores had higher increases in the incidence rate between 14 April and 15 May. States with medium human development index (HDI) showed higher incidence increases from 14 April to 15 May, being seven of them with ratios in the range from 27.49 to 63.73 times.

**Conclusion:** Spreading of COVID-19 in Brazil differs across both regions and federative units, being influenced by different socioeconomic contexts.

## Introduction

This year 2020 will go down in world history due to the coronavirus disease of 2019 (COVID-19) pandemic. The first suspected case of COVID-19 was reported in Wuhan, the capital of Hubei Province, China, on 1 December 2019 [1-3]. The peak of the epidemic in China occurred on 19 February 2020, and a month later the number of deaths in China reached 3,242 out of a total of 81,174 confirmed cases [4].

In a health crisis unprecedented in recent history, cases of severe acute respiratory syndrome by coronavirus 2 (SARS-CoV-2) crossed the country’s borders and rapidly spread across the world, leading to the official declaration of a global pandemic and public health emergency by the World Health Organization (WHO) in 11 March 2020 [5, 6].

Person-to-person transmission of SARS-CoV-2 from asymptomatic patients was first documented in Germany and Vietnam clusters during early February 2020 [7, 8]. However, Wu and McGoogan (2020) described that 81% of initial patients notified in China experienced only mild disease, which could have contributed to an asymptomatic transmission [9]. In addition, recent reports have pointed out that virus incubation may be longer than the 14 days period used by the WHO to guide quarantine policies [10, 11]. Taken together, these factors are undoubtedly contributing to the rapid spread of SARS-CoV-2 over the world [12-14].

Since early March 2020, a rapid increase in the number of cases in Italy, Spain, France, Germany and Iran has been observed. According to data from WHO (situation report 123) on 22 May 2020, most reported cases occurred in the United States of America (1,525,186 cases with 91,527 deaths), the Russian Federation (326,448 cases with 3,249 deaths) followed by Brazil in third position [15].

In Brazil, the first confirmed case of COVID-19 was notified in São Paulo state on 26 February, while the first death was registered on 17 March 2020 in Rio de Janeiro state. Since then, the incidence of COVID-19 has increased very rapidly in the country, reaching a total of 330,890 confirmed cases with 21,048 deaths, by the cut-off date for data compilation in this study, 22 May 2020 [16].

In this context, epidemiological studies conducted early in the course of an epidemic are important to guide public health decision-making. Monitoring numbers of COVID-19 cases, deaths, as well as regional differences in incidence, is critical for understanding community risks and making decisions about strategic allocation of health care resources and the control of social distancing during the disease outbreak. Thus, the aim of this study was to document and monitor the increase of COVID-19 cases across each federative unit in Brazil, by following the progression of notified cases from inception to 15 May 2020, as well as their determinants and health indicators.

## Material and Methods

### Study Design

This is an observational study that tracked data on notified cases of COVID-19 in Brazil, from inception to 15 May 2020.

### Data collection

Data on the numbers of both COVID-19 cases and deaths from the disease were obtained from the official website of the Ministry of Health of Brazil, from 14 April 2020 onwards [16]. When the data from the Brazilian government website were consulted, care was taken to select the date of the first notification of COVID-19 and the data on 14 April. From that date, a daily follow-up of cases was conducted in each federative unit until 15 May 2020.

Sociodemographic data (population and number of elderly people) by cities and states were estimated by the Brazilian Institute of Geography and Statistics (IBGE) based on the index-date of 1 July 2019, from the last census conducted in Brazil in 2010 [17]. Per capita income was extracted from the demographic census conducted in 2010 [18].

Data on health determinants, such as percentage of inadequate water supply and sewerage, and the number of hospital beds and intensive treatment units, were obtained from DATASUS, the IT Department of the Brazilian Unified Health System, and IBGE [18, 19]. In addition, social isolation rates were obtained from Inloco website [20].

The incidence rate of COVID-19 was obtained based on the estimated population of each federative unit in 2019 and it was presented as cases per 100,000. The case fatality was calculated and presented as a percentage.

The Human Development Index (HDI) scores were obtained from the Brazilian Atlas of Human Development, produced by United Nations Development Program (UNDP), Institute of Applied Economic Research (IPEA) and João Pinheiro Foundation [21]. HDI is a development indicator that aggregates economic data from the per capita gross domestic product with other data related to education [adult literacy rate and schooling (combined primary, secondary, and higher education)] and health (mean life expectancy at birth). HDI scores can range from 0 to 1, and the 27 Brazilian federative units are grouped into four development categories, depending on their HDI score: very high for HDI ≥ 0.800, high from 0.700 to 0.799, average from 0.550 to 0.699 and low when HDI is below 0.550 [21].

The Social Vulnerability Index (SVI) was developed by IPEA. SVI scores were calculated based on the 2010 Brazil census variables. This tool is particularly useful for emergency response planners and public health authorities, as it can identify and map communities most likely to need support before, during, and after a hazardous event. SVI scores can vary from 0 to 1, with higher values associated to higher social vulnerability. SVI scores for each municipality were classified in five levels: very low, from 0 to 0.200; low, from 0.201 to 0.300; medium, from 0.301 to 0.400; high, from 0.401 to 0.500; and very high, above 0.501 to 1 [22].

In addition, data were organized and made available at a geospatial data platform called PIE-COVID, which has numerous georeferenced variables of all cases of COVID-19 in Brazilian cities and states [23].

### Ethics

In this study, only public domain data was used, without identifying personal data. According to the Resolution number 510/2016 from Brazilian legislation of the National Health Council *(Conselho Nacional de Saúde (CNS))*, no ethical approval is required under Ethics Committee on Research *(Comitê de Ética em Pesquisa* (CEP) and National Ethics Commission *(Comissão Nacional de Ética (Conep))* [24].

### Statistical analyses

Statistical analyses were performed using SPSS version 22.0 (IBM, USA) and GraphPad Prism™ version 5.04 (GraphPad Software, Inc., USA).

Spearman’s correlations between COVID-19 incidence on 31 April and SVI, and between incidence ratio (15 May/14 April) and SVI, were calculated for each of the 27 federative units.

Incidence rates of COVID-19 on 15 May were divided into two groups, according to median rate (cut-off point, >100). Water supply and sewerage inadequacy rates (%) were also categorized in two groups, according to median rate (cut-off point, ≥9.0%).

The increase in incidence rate over the course of a month was obtained by the ratio between incidence rates registered on 15 May and 14 April 2020. In addition, increases in incidence rates were classified into quartiles (1^st^ quartile, 2.83 to 5.94; 2^nd^ quartile, 5.95 to 13.38; 3^rd^ quartile, 13.39 to 27.48; and 4^th^ quartile, 27.49 to 63.73).

The chi-square test or the likelihood ratio were used to analyse frequencies of categorical variables, with a significance level of α = 0.05.

## Results

The first case of COVID-19 in Brazil was reported in São Paulo city on 26 February, a person who had returned from a trip to Italy some days before. On this date, Brazil became the first country to report a case of COVID-19 in Latin America.

Among the capitals of Brazilian federative units, 25 out of 27 had cases of COVID-19 in March. Only the capitals of the states of Piauí (Teresina) and Tocantins (Palmas) reported cases in April. In Piauí, two cities had cases in March (Parnaíba and São José do Divino), and in Tocantins only Araguaína had a case on 27 March 2020.

In this study, we decided to present data according to the five politic regions of Brazil: North, Northeast, Southeast, South and Midwest.

North region consists of seven states: Rondônia (RO), Acre (AC), Amazonas (AM), Roraima (RR), Pará (PA), Amapá (AP), and Tocantins (TO). The first case of COVID-19 recorded in this region was on 15 March 2020 in AM, the state with the worst social vulnerability index (SVI) and one of the worst Human Development Index (HDI) scores among North states.

Furthermore, AM together with the small state of AP registered on 30 April 2020 the highest incidence of COVID-19 in the North. On 15 May 2020, the incidence rate of COVID-19 in these states (AM, 443.76; and AP, 429.21) was more than three times that of São Paulo (127.13), Federal District (125.59), and Rio de Janeiro (115.77), Fig 1. The North region had 40,473 cases and 2,762 deaths, ranking third among all five regions. Information about population, health determinants and indicators and about notification of COVID-19 of North region from the beginning of pandemic until 15 May 2020 is shown in Table 1 and Fig 1 and 2.

**Figure 1.**
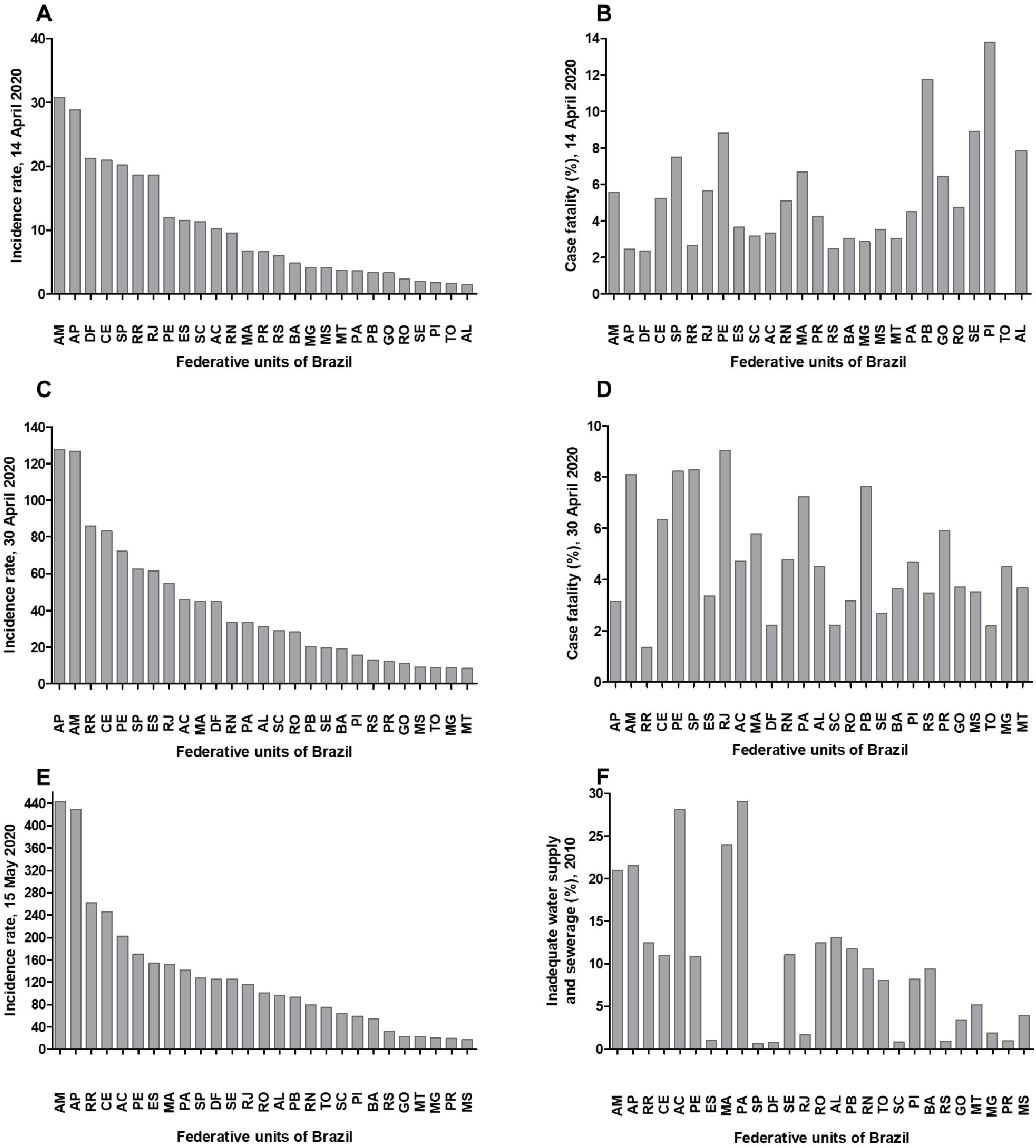
**Incidence rate of COVID-19 (cases per 100,000) and case fatality (%) in Brazilian federative units on 14 (A and B) and 30 (C and D) April 2020; incidence rate of COVID-19 on 15 May 2020 (E); and inadequate water supply and sewerage rates (F), 2010. Figure F shows federative units in decreasing order of COVID-19 incidence, according to data from 15 May 2020**. AC: Acre, AL: Alagoas, AP: Amapá, AM: Amazonas, BA: Bahia, CE: Ceará, DF: Distrito Federal, ES: Espírito Santos, GO: Goiás, MA: Maranhão, MT: Mato Grosso, MS: Mato Grosso do Sul, MG: Minas Gerais, PA: Pará, PB: Paraíba, PR: Paraná, PE: Pernambuco, PI: Piauí, RJ: Rio de Janeiro, RN: Rio Grande do Norte, RS: Rio Grande do Sul, RO: Rondônia, RR: Roraima, SC: Santa Catarina, SP: São Paulo, SE: Sergipe, TO: Tocantins.

**Table 1.** Notification of COVID-19 in the North and Northeast r 213 egions of Brazil until 15 May 2020.

| NORTH | RO | AC | AM | RR | PA | AP | TO | Total |  |  |
| --- | --- | --- | --- | --- | --- | --- | --- | --- | --- | --- |
| Population<sup>1</sup> | 1,777,225<br>9.64% | 881,935<br>4.79% | 4,144,597<br>22.49% | 605,761<br>3.29% | 8,602,865<br>46.68% | 845,731<br>4.59% | 1,572,866<br>8.53% | 18,430,980<br>100% |  |  |
| Elderly (≥65 y)<sup>1</sup> | 107,581 | 45,712 | 202,464 | 25,770 | 506,893 | 36,085 | 112,964 | 1,037,469 |  |  |
| Total number of cities | 52 | 22 | 62 | 15 | 144 | 16 | 139 | 450 |  |  |
| Per capita income (R\$) | 670.82 | 522.15 | 539.80 | 605.59 | 446.76 | 598.98 | 586.62 | | | |
| Inadequate water supply and sewerage (%) | 12.43 | 28.09 | 20.98 | 12.44 | 29.05 | 21.51 | 8.07 |  |  |  |
| SVI | 0.319 | 0.443 | 0.488 | 0.366 | 0.469 | 0.404 | 0.336 |  |  |  |
| HDI | 0.690 | 0.663 | 0.674 | 0.707 | 0.646 | 0.708 | 0.699 |  |  |  |
| Hospital Beds in SUS | 3,360 | 1,350 | 4,914 | 1,064 | 10,240 | 953 | 2,327 | 24,208 |  |  |
| ICU beds in SUS | 287 | 133 | 691 | 87 | 1,081 | 91 | 235 | 2,605 |  |  |
| Total hospital beds | 4,286 | 1,484 | 5,700 | 1,113 | 13,483 | 1,098 | 3,072 | 30,256 |  |  |
| Population/hospital beds | 415 | 594 | 727 | 535 | 638 | 770 | 512 | 609 |  |  |
| Date of 1<sup>st</sup> case | 20.03.20 | 19.03.20 | 15.03.20 | 22.03.20 | 19.03.20 | 20.03.20 | 19.03.20 | 15.03.20 |  |  |
| Total cases, March | 8 | 42 | 175 | 16 | 32 | 10 | 11 | 294 |  |  |
| Deaths, March | 1 | 0 | 3 | 0 | 0 | 0 | 0 | 4 |  |  |
| Social isolation (%) in 14 April 2020 | 37.89 | 39.70 | 47.90 | 38.06 | 44.78 | 47.31 | 35.54 |  |  |  |
| Cases until 14 April 2020 | 42 | 90 | 1,276 | 113 | 311 | 244 | 26 | 2,102 |  |  |
| Deaths until 14 April 2020 | 2 | 3 | 71 | 3 | 14 | 6 | 0 | 99 |  |  |
| Incidence on 14 April 2020* | 2.36 | 10.20 | 30.79 | 18.65 | 3.62 | 28.85 | 1.65 | 11.40 |  |  |
| Social isolation (%) in 30 April 2020 | 46.8 | 50.9 | 52.6 | 48.2 | 57.9 | 60.0 | 48.3 |  |  |  |
| Cases until 30 April 2020 | 502 | 404 | 5,254 | 519 | 2,876 | 1,080 | 137 | 10,772 |  |  |
| Deaths until 30 April 2020 | 16 | 19 | 425 | 7 | 208 | 34 | 3 | 712 |  |  |
| Incidence on 30 April 2020* | 28.25 | 45.81 | 126.77 | 85.68 | 33.43 | 127.7 | 8.71 | 58.45 |  |  |
| Cases until 15 May 2020 | 1,789 | 1,785 | 18,392 | 1,589 | 12,109 | 3,630 | 1,179 | 40,473 |  |  |
| Deaths until 15 May 2020 | 62 | 57 | 1,331 | 40 | 1,145 | 103 | 24 | 2,762 |  |  |
| Incidence on 15 May 2020 | 100.66 | 202.40 | 443.76 | 262.31 | 140.76 | 429.21 | 74.96 | 219.59 |  |  |
| NORTHEAST | MA | PI | CE | RN | PB | PE | AL | SE | BA | Total |
| Population<sup>1</sup> | 7,075,181<br>12.40% | 3,273,227<br>5.74% | 9,132,078<br>16.00% | 3,506,853<br>6.14% | 4,018,127<br>7.04% | 9,557,071<br>16.75% | 3,337,357<br>5.85% | 2,298,696<br>4.03% | 14,873,064<br>26.06% | 57,071,654<br>100% |
| Elderly (≥65 y)<sup>1</sup> | 503,654 | 285,899 | 816,935 | 315,584 | 393,136 | 845,808 | 254,662 | 172,546 | 1,335,027 | **4,923,251** |
| Total number of cities | 217 | 224 | 184 | 167 | 223 | 185 | 102 | 75 | 417 | **1,794** |
| Per capita income (R\$) | 360.34 | 416.93 | 460.63 | 545.42 | 474.94 | 525.64 | 432.56 | 523.53 | 496.73 | |
| Inadequate water supply and sewerage (%) | 23.99 | 8.15 | 10.99 | 9.4 | 11.75 | 10.83 | 13.07 | 11.02 | 9.35 |  |
| SVI | 0.521 | 0.403 | 0.378 | 0.349 | 0.385 | 0.414 | 0.461 | 0.393 | 0.403 |  |
| IDH | 0.639 | 0.646 | 0.682 | 0.684 | 0.658 | 0.673 | 0.631 | 0.665 | 0.660 |  |
| Hospital Beds in SUS | 12,204 | 6,470 | 14,350 | 5,958 | 6,425 | 15,644 | 4,807 | 2,376 | 22,947 | **91,181** |
| ICU beds in SUS | 784 | 392 | 1,243 | 473 | 671 | 1,404 | 488 | 348 | 1,772 | **7,575** |
| Total hospital beds | 13,778 | 7,460 | 18,510 | 7,250 | 8,147 | 20,945 | 5,891 | 3,212 | 28,960 | **114,153** |
| Population/hospital beds | 514 | 439 | 493 | 484 | 493 | 456 | 567 | 716 | 514 | **500** |
| Date of 1<sup>st</sup> case | 21.03.20 | 20.03.20 | 17.03.20 | 13.03.20 | 19.03.20 | 12.03.20 | 08.03.20 | 15.03.20 | 06.03.20 | **06.03.20** |
| Total cases March | 31 | 18 | 390 | 82 | 17 | 87 | 18 | 19 | 213 | **875** |
| Deaths March | 1 | 4 | 7 | 1 | 0 | 6 | 1 | 0 | 2 | **22** |
| Social isolation (%) in 14 April 2020 | 47.54 | 47.95 | 49.97 | 85.86 | 46.81 | 50.60 | 44.25 | 47.00 | 46.30 |  |
| Cases until 14 April 2020 | 479 | 58 | 1911 | 333 | 136 | 1146 | 51 | 45 | 722 | **4,881** |
| Deaths until 14 April 2020 | 32 | 8 | 100 | 17 | 16 | 101 | 4 | 4 | 22 | **304** |
| Incidence on 14 April 2020* | 6.77 | 1.77 | 20.93 | 9.50 | 3.38 | 11.99 | 1.53 | 1.96 | 4.85 | **8.55** |
| Social isolation (%) in 30 April 2020 | 59.6 | 59.0 | 57.9 | 55.7 | 55.3 | 58.5 | - | 53.2 | 52.9 |  |
| Cases until 30 April 2020 | 3,190 | 513 | 7,606 | 1,177 | 814 | 6,876 | 1,044 | 447 | 2,851 | **24,518** |
| Deaths until 30 April 2020 | 184 | 24 | 482 | 56 | 62 | 565 | 47 | 12 | 104 | **1,536** |
| Incidence on 30 April 2020* | 45.09 | 15.67 | 83.29 | 33.56 | 20.26 | 71.95 | 31.28 | 19.45 | 19.17 | **42.96** |
| Cases until 15 May 2020 | 10,739 | 1,905 | 22,490 | 2,786 | 3,739 | 16,209 | 3,212 | 2,868 | 8,128 | **72,076** |
| Deaths until 15 May 2020 | 496 | 60 | 1,476 | 122 | 170 | 1,381 | 187 | 50 | 281 | **4,223** |
| Incidence on 15 May 2020 | 151.78 | 58.2 | 246.27 | 79.44 | 93.05 | 169.60 | 96.24 | 124.77 | 54.65 | **126.29** |

<sup>1</sup> Data estimated by the IBGE for the year 2019. All indicators were based on IBGE 2010 data. Federative units in the North region of Brazil are RO, Rondônia; AC, Acre; AM, Amazonas; RR, Roraima; PA, Pará; AP, Amapá; and TO, Tocantins; and in the Northeast region of Brazil are MA, Maranhão; PI, Piauí; CE, Ceará; RN, Rio Grande do Norte; PB, Paraíba; PE, Pernambuco; AL, Alagoas; SE, Sergipe; and BA, Bahia. SVI, Social vulnerability index; HDI, Human Development index; SUS, Brazilian Unified Public Health System; ICU, Intensive Care United. \*Incidence was shown by cases per 100,000 people, according to population data estimated by the IBGE for the year 2019.

**Figure 2.**
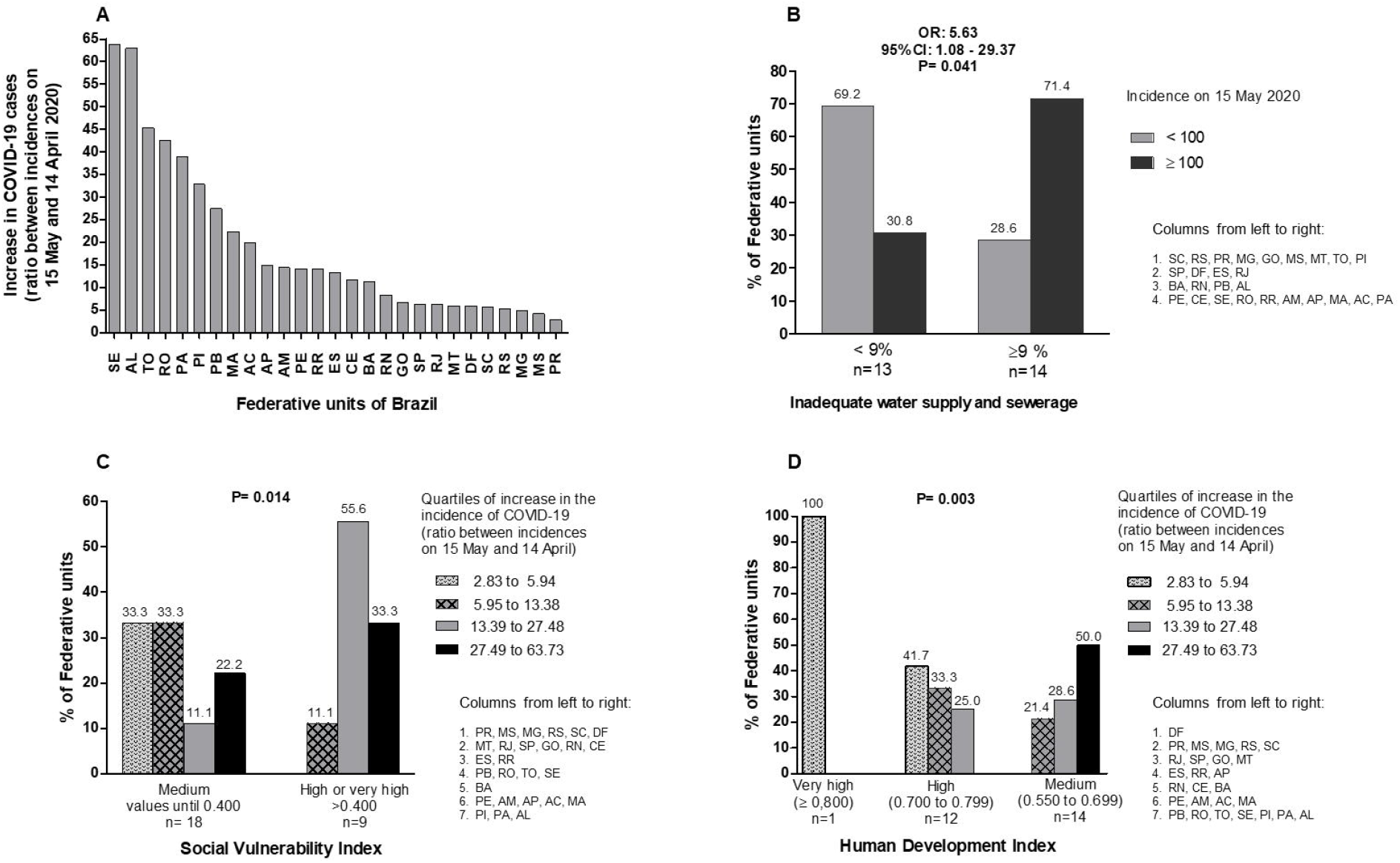
**Data on the incidence of COVID-19 in Brazil until 15 May 2020. (A) Increase in cases over a period of 31 days, using the ratio between incidences per 100,000 on 15 May and 14 April 2020. (B) Association between the incidence of COVID-19 on 15 May (classified into two groups, according to the median) and percentage of inadequate water supply and sewerage (classified into two groups, according to the median). (C) Association between quartiles of increase in the incidence of COVID-19 (incidence ratio) and scores on the Social Vulnerability Index (SVI) (classified into SVI scores ≤ “mean” and SVI scores “high” or “very high”). (D) Association between quartiles of increase in the incidence of COVID-19 (incidence ratio) and scores on the Human Development Index (classified into “very high” (≥ 0.800), “high” (0.700 to 0.799) and “medium” (0.550 to 0.699)). B. Chi-square test. C and D. Likelihood ratio**. AC, Acre; AL, Alagoas; AP, Amapá; AM, Amazonas; BA, Bahia; CE, Ceará; DF, Distrito Federal; ES, Espírito Santos; GO, Goiás; MA, Maranhão; MT, Mato Grosso; MS, Mato Grosso do Sul; MG, Minas Gerais; PA, Pará; PB, Paraíba; PR, Paraná; PE, Pernambuco; PI, Piauí; RJ, Rio de Janeiro; RN, Rio Grande do Norte; RS, Rio Grande do Sul; RO, Rondônia; RR, Roraima; SC, Santa Catarina; SP, São Paulo; SE, Sergipe; TO, Tocantins.

Regarding the Northeast region, it is composed of nine states, namely: Maranhão (MA), Piauí (PI), Ceará (CE), Rio Grande do Norte (RN), Paraíba (PB), Pernambuco (PE), Alagoas (AL), Sergipe (SE), and Bahia (BA). The first case in this region was described on 6 March 2020, in BA. As of 15 May 2020, 72,076 cases and 4,223 deaths were reported in Northeast, with CE and PE showing both the highest incidence rates and deaths numbers. Until the last data collection for this study, the cities in Northeast with the highest numbers of confirmed cases and deaths were Fortaleza (CE), Recife (PE), and São Luis (MA). Detailed information on population, health determinants and indicators, and on notification of COVID-19 cases in the Northeast region from the first case reported until 15 May 2020 are shown in Table 1.

Southeast region includes the states of Minas Gerais (MG), Espírito Santo (ES), Rio de Janeiro (RJ) and São Paulo (SP), with a large number of municipalities (1,668). Both the first confirmed case and death by COVID-19 registered in this region were also the first ones reported in the country. On 15 May 2020, the southeastern region ranks first in the number of cases (88,759) and deaths (7,345) in Brazil.

In this region, São Paulo state shows the highest number of cases (58,378) with 4,501 deaths until 15 May 2020. In the city of São Paulo alone, 33,992 cases were confirmed with 2,674 deaths, making it the most affected municipality in the Southeast, followed by Rio de Janeiro, with 11.715 cases and 1,631 deaths on 15 May. In MG, the city with the highest number of cases and deaths, Belo Horizonte, had 1,088 confirmed cases and 29 deaths, while in ES the largest number of cases occurred in Vila Velha, a city very close to the capital (Vitória), with 1,305 and 43 deaths. Complete data on population, health determinants and indicators, as well as on notification of COVID-19 cases in the Southeast region, from the beginning of pandemic until 15 May 2020, are shown in Table 2.

The South region comprises the states of Paraná (PR), Santa Catarina (SC) and Rio Grande do Sul (RS). The first confirmed patient with COVID-19 in the region was in the city of Campo Bom, RS. In this region, the state of SC stands out with the highest incidence in the South on 14 April and the highest increase until 15 May. Information about population, health determinants and indicators and about notification of COVID-19 cases in the South region, from the first registered case until 15 May 2020, are shown in Table 3.

Finally, the Midwest region is composed of the states of Mato Grosso (MT), Mato Grosso do Sul (MS), Goiás (GO) and the Federal District (DF). The first confirmed case in the region occurred on 7 March 2020, in DF. On 15 May 2020, DF had 3,787 confirmed cases and 55 deaths. Complete data for this region is also showed in Table 3 and Fig 1 and 2.

**Table 2.** Notification of COVID-19 in the Southeast 280 region of Brazil until 15 May 2020.

| **SOUTHEAST** | **MG** | **ES** | **RJ** | **SP** | **Total** |
| --- | --- | --- | --- | --- | --- |
| Population<sup>1</sup> | 21,168,791 | 4,018,650 | 17,264,943 | 45,919,049 | **88,371,433** |
|  | 23.95% | 4.55% | 19.54% | 51.96% | **100%** |
| Elderly (≥65 y)<sup>1</sup> | 2,288,973 | 382,976 | 1,998,280 | 4,792,036 | **9,462,265** |
| Total number of cities | 853 | 78 | 92 | 645 | **1,668** |
| Per capita income (R\$) | 749.69 | 815.43 | 1039.3 | 1084.46 | |
| Inadequate water supply and sewerage (%) | 1.84 | 0.99 | 1.67 | 0.6 |  |
| SVI | 0.282 | 0.274 | 0.323 | 0.297 |  |
| HDI | 0.731 | 0.740 | 0.761 | 0.783 |  |
| Hospital beds in SUS | 26,896 | 5,100 | 21,108 | 52,302 | **105,406** |
| ICU beds in SUS | 3,478 | 641 | 2,612 | 7,331 | **14,062** |
| Total hospital beds | 40,675 | 7,929 | 32,736 | 90,146 | **171,486** |
| Population/hospital beds | 520 | 507 | 527 | 509 | **515** |
| Date of 1<sup>st</sup> case | 08.03.20 | 06.03.20 | 05.03.20 | 26.02.20 | **26.02.20** |
| Total cases, March | 275 | 84 | 708 | 2339 | **3,406** |
| Deaths, March | 2 | 0 | 23 | 136 | **161** |
| Social isolation (%) in 14 April 2020 | 43.35 | 44.49 | 48.59 | 46.8 |  |
| Cases until 14 April 2020 | 870 | 463 | 3,220 | 9,263 | **4,553** |
| Death until 14 April 2020 | 25 | 17 | 182 | 695 | **224** |
| Incidence on 14 April 2020* | 4.11 | 11.52 | 18.65 | 20.17 | **5.15** |
| Social isolation (%) in 30 April 2020 | 51.0 | 51.3 | 54.4 | 51.4 |  |
| Cases until 30 April 2020 | 1,827 | 2,465 | 9,453 | 28,698 | **42,443** |
| Death until 30 April 2020 | 82 | 83 | 854 | 2,375 | **3,394** |
| Incidence on 30 April 2020* | 8.63 | 61.34 | 54.75 | 62.5 | **48.03** |
| Cases until 15 May 2020 | 4,196 | 6,198 | 19,987 | 58,378 | **88,759** |
| Death until 15 May 2020 | 146 | 260 | 2,438 | 4,501 | **7,345** |
| Incidence on 15 May 2020 | 19.82 | 154.23 | 115.77 | 127.13 | **100.44** |

<sup>1</sup> Data estimated by the IBGE for the year 2019. All indicators were based on IBGE 2010 data. Federative units in the Southeast region of Brazil are MG, Minas Gerais; ES, Espírito Santo; RJ, Rio de Janeiro; and SP, São Paulo. SVI, Social vulnerability index; HDI, Human Development index; SUS, Brazilian Unified Public Health System; ICU, Intensive Care Unit. \*Incidence was shown as cases per 100,000 people, according to population data estimated by the IBGE for the year 2019.

**Table 3.**
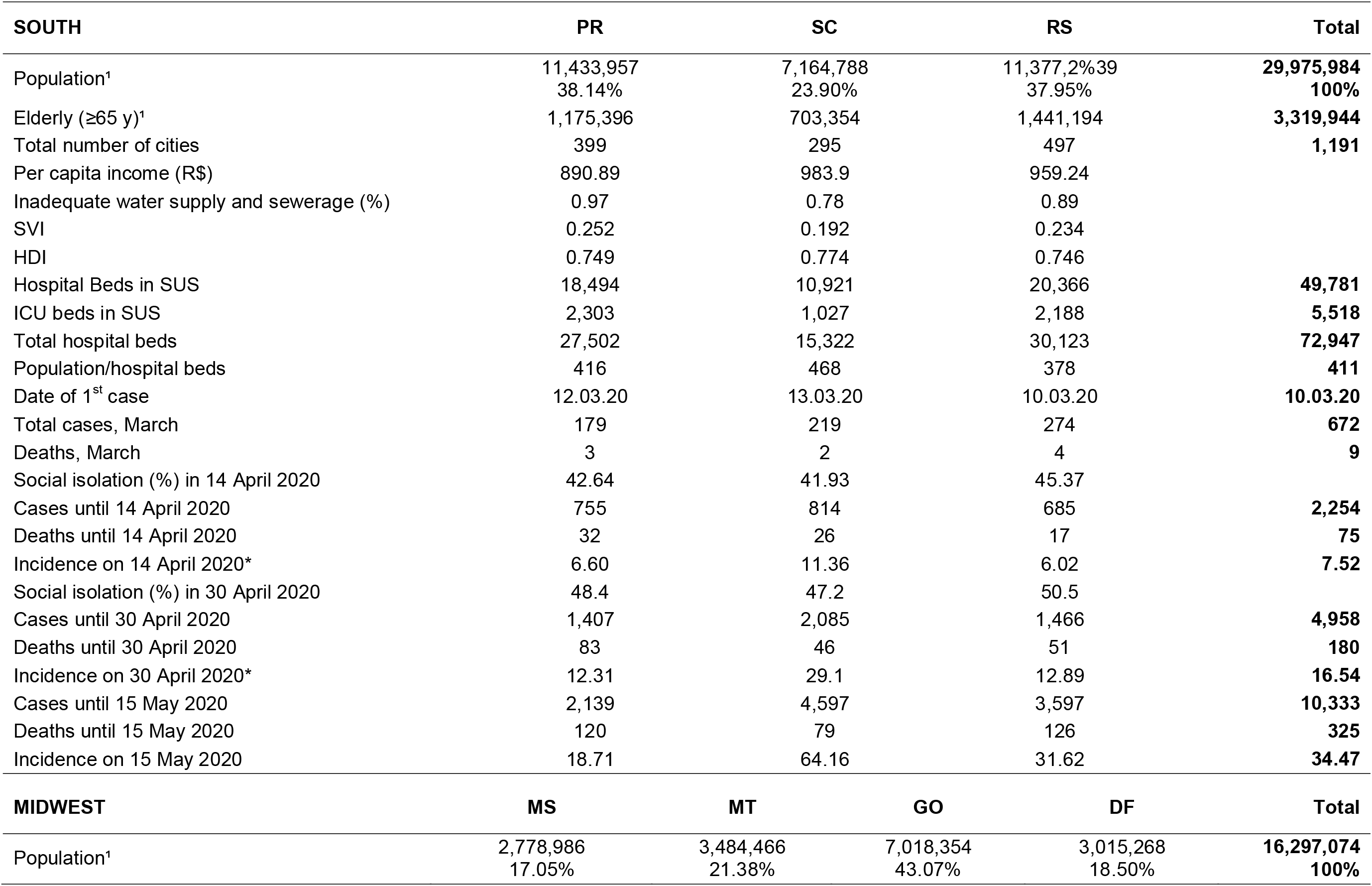
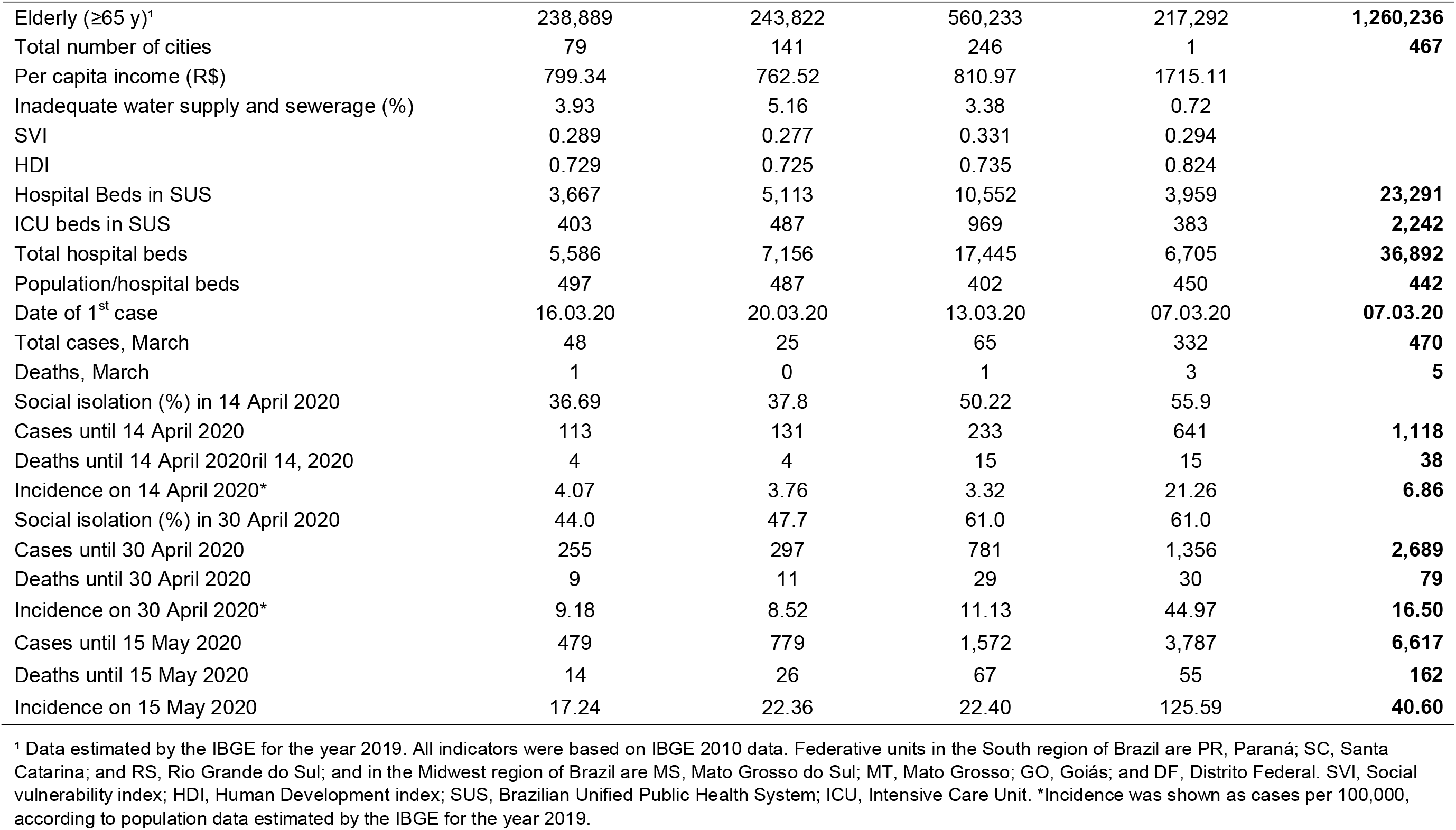
Notification of COVID-19 in the 285 South and Midwest regions of Brazil until 15 May 2020.

| SOUTH | PR | SC | RS | Total |  |
| --- | --- | --- | --- | --- | --- |
| Population¹ | 11,433,957<br>38.14% | 7,164,788<br>23.90% | 11,377,2%39<br>37.95% | 29,975,984<br>100% |  |
| Elderly (≥65 y)¹ | 1,175,396 | 703,354 | 1,441,194 | 3,319,944 |  |
| Total number of cities | 399 | 295 | 497 | 1,191 |  |
| Per capita income (R\$) | 890.89 | 983.9 | 959.24 | | |
| Inadequate water supply and sewerage (%) | 0.97 | 0.78 | 0.89 |  |  |
| SVI | 0.252 | 0.192 | 0.234 |  |  |
| HDI | 0.749 | 0.774 | 0.746 |  |  |
| Hospital Beds in SUS | 18,494 | 10,921 | 20,366 | 49,781 |  |
| ICU beds in SUS | 2,303 | 1,027 | 2,188 | 5,518 |  |
| Total hospital beds | 27,502 | 15,322 | 30,123 | 72,947 |  |
| Population/hospital beds | 416 | 468 | 378 | 411 |  |
| Date of 1<sup>st</sup> case | 12.03.20 | 13.03.20 | 10.03.20 | 10.03.20 |  |
| Total cases, March | 179 | 219 | 274 | 672 |  |
| Deaths, March | 3 | 2 | 4 | 9 |  |
| Social isolation (%) in 14 April 2020 | 42.64 | 41.93 | 45.37 |  |  |
| Cases until 14 April 2020 | 755 | 814 | 685 | 2,254 |  |
| Deaths until 14 April 2020 | 32 | 26 | 17 | 75 |  |
| Incidence on 14 April 2020* | 6.60 | 11.36 | 6.02 | 7.52 |  |
| Social isolation (%) in 30 April 2020 | 48.4 | 47.2 | 50.5 |  |  |
| Cases until 30 April 2020 | 1,407 | 2,085 | 1,466 | 4,958 |  |
| Deaths until 30 April 2020 | 83 | 46 | 51 | 180 |  |
| Incidence on 30 April 2020* | 12.31 | 29.1 | 12.89 | 16.54 |  |
| Cases until 15 May 2020 | 2,139 | 4,597 | 3,597 | 10,333 |  |
| Deaths until 15 May 2020 | 120 | 79 | 126 | 325 |  |
| Incidence on 15 May 2020 | 18.71 | 64.16 | 31.62 | 34.47 |  |
| MIDWEST | MS | MT | GO | DF | Total |
| Population¹ | 2,778,986<br>17.05% | 3,484,466<br>21.38% | 7,018,354<br>43.07% | 3,015,268<br>18.50% | 16,297,074<br>100% |
| Elderly (≥65 y)<sup>1</sup> | 238,889 | 243,822 | 560,233 | 217,292 | **1,260,236** |
| Total number of cities | 79 | 141 | 246 | 1 | **467** |
| Per capita income (R\$) | 799.34 | 762.52 | 810.97 | 1715.11 | |
| Inadequate water supply and sewerage (%) | 3.93 | 5.16 | 3.38 | 0.72 |  |
| SVI | 0.289 | 0.277 | 0.331 | 0.294 |  |
| HDI | 0.729 | 0.725 | 0.735 | 0.824 |  |
| Hospital Beds in SUS | 3,667 | 5,113 | 10,552 | 3,959 | **23,291** |
| ICU beds in SUS | 403 | 487 | 969 | 383 | **2,242** |
| Total hospital beds | 5,586 | 7,156 | 17,445 | 6,705 | **36,892** |
| Population/hospital beds | 497 | 487 | 402 | 450 | **442** |
| Date of 1<sup>st</sup> case | 16.03.20 | 20.03.20 | 13.03.20 | 07.03.20 | **07.03.20** |
| Total cases, March | 48 | 25 | 65 | 332 | **470** |
| Deaths, March | 1 | 0 | 1 | 3 | **5** |
| Social isolation (%) in 14 April 2020 | 36.69 | 37.8 | 50.22 | 55.9 |  |
| Cases until 14 April 2020 | 113 | 131 | 233 | 641 | **1,118** |
| Deaths until 14 April 2020ril 14, 2020 | 4 | 4 | 15 | 15 | **38** |
| Incidence on 14 April 2020* | 4.07 | 3.76 | 3.32 | 21.26 | **6.86** |
| Social isolation (%) in 30 April 2020 | 44.0 | 47.7 | 61.0 | 61.0 |  |
| Cases until 30 April 2020 | 255 | 297 | 781 | 1,356 | **2,689** |
| Deaths until 30 April 2020 | 9 | 11 | 29 | 30 | **79** |
| Incidence on 30 April 2020* | 9.18 | 8.52 | 11.13 | 44.97 | **16.50** |
| Cases until 15 May 2020 | 479 | 779 | 1,572 | 3,787 | **6,617** |
| Deaths until 15 May 2020 | 14 | 26 | 67 | 55 | **162** |
| Incidence on 15 May 2020 | 17.24 | 22.36 | 22.40 | 125.59 | **40.60** |

<sup>1</sup> Data estimated by the IBGE for the year 2019. All indicators were based on IBGE 2010 data. Federative units in the South region of Brazil are PR, Paraná; SC, Santa Catarina; and RS, Rio Grande do Sul; and in the Midwest region of Brazil are MS, Mato Grosso do Sul; MT, Mato Grosso; GO, Goiás; and DF, Distrito Federal. SVI, Social vulnerability index; HDI, Human Development index; SUS, Brazilian Unified Public Health System; ICU, Intensive Care Unit. \*Incidence was shown as cases per 100,000, according to population data estimated by the IBGE for the year 2019.

Incidence rates and case fatality rates (%) in three time points (14 and 30 April, and 15 May), ranked in decreasing order, as well as SVI scores and inadequate water supply and sewerage rates (%), by federative unit, are demonstrated in Fig 1. Regarding incidence rates (cases per 100,000), the top five federative units in 14 April 2020, in decreasing order, were AM and AP (North), DF (Midwest), CE (Northeast) and SP (Southeast), all with more than 20 cases per 100,000, Fig 1A. In 30 April 2020, the states with highest incidence rates were AP, AM and RR (North), CE and PE (Northeast), all with incidence rates above 70 cases per 100,000. As for fatality rates, considering all 27 states in the country, RJ and SP had the highest scores, followed by PE, AM and PB. It is important to highlight that, despite PB is just the 17^th^ of 27 states in terms of incidence, it appears as the fifth when lethality is measured (Fig 1C). In the last evaluation, the top of five federative units in 15 May 2020, in decreasing order, were AM (incidence: 443.76), AP (429.21) and RR (262.31) (North), CE with incidence 246.27 (Northeast) and AC (246.27) (North). Fourteen states had incidences greater than 150 per 100.00 on that date Fig 1E.

The increase in cases over the course of a month, based on the ratio between incidences per 100,000 on 15 May and 14 April 2020, is shown in Fig 2A. Considering the increasing in incidence rates of COVID-19 cases during this period, the states with the highest levels, in decreasing order, were SE, AL, TO, PA and PI.

When federative units were stratified into two groups according to the rate of inadequate water supply and sewerage (< 9% or ≥ 9%), those with rates >9% were 5.63 times more likely to have an incidence greater than 30 cases/100,000 (P=0.041), as shown in Fig 2B.

Stratifying increases in cases over this one-month period into quartiles revealed how SVI and HDI scores were strongly associated with higher values of incidence ratio in North and Northeast states (Fig 2C and D).

The SVI data for the 27 federative units were moderately and directly correlated with the incidence of cases on 15 May (rh_0_ = 0.575 and P =0.002). There was a strong and direct correlation between increases in the incidence rate during the one-month period covered in the study and SVI scores (rh_0_ = 0.727 and P <0.001).

Finally, updated data on numbers of cases, incidence rates, number of deaths, and case-fatality, until 22 May 2020 is presented in Fig 3. Data about incidence rate of COVID-19 on 22 May 2020 could be found in S1 Fig.

**Figure 3.**
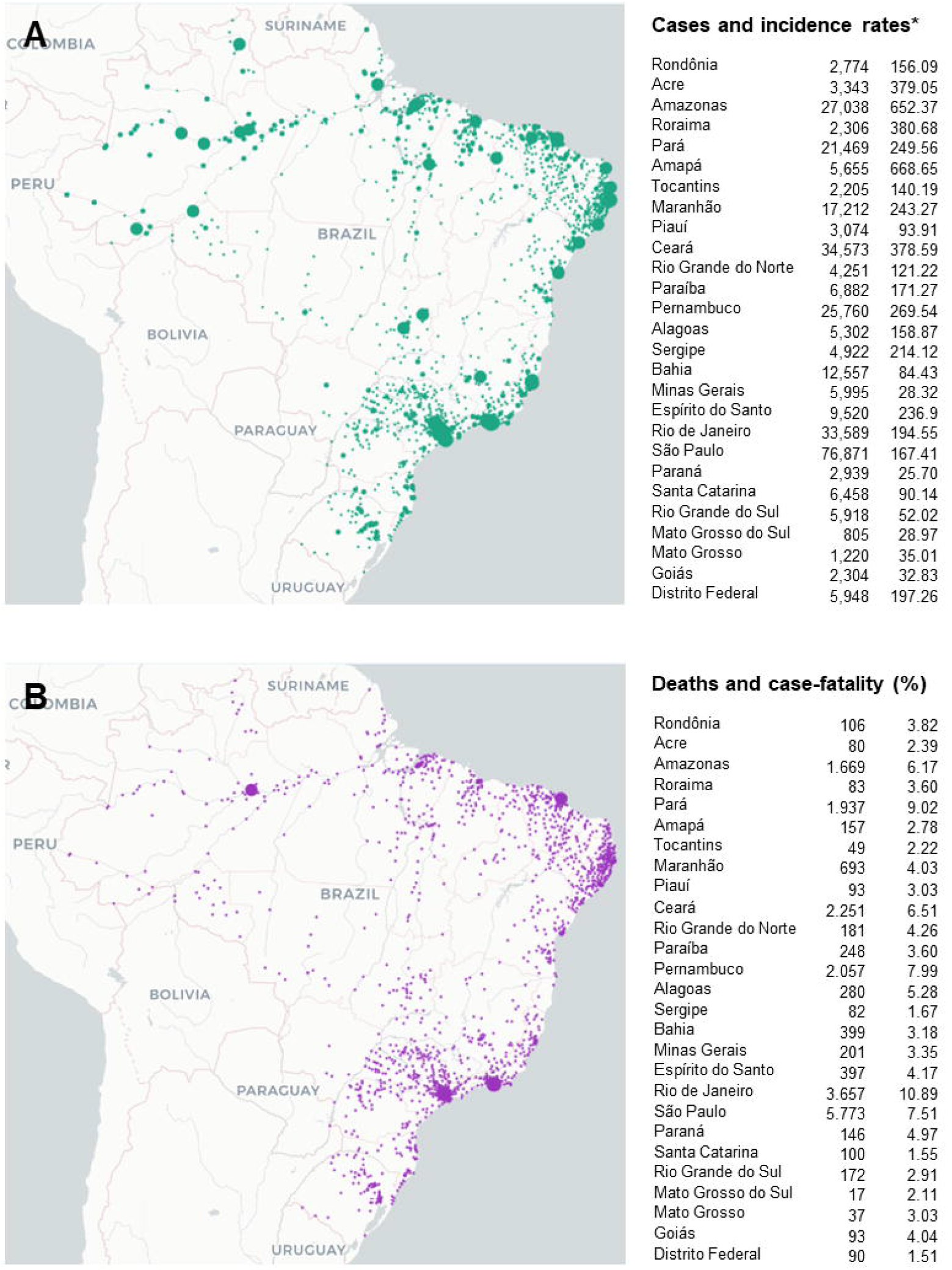
**Distribution of cases and incidence rates (A), deaths and case-fatality (B) in the 27 federative units of Brazil on 22 May 2020 [16]**. * Incidence rates were calculated according to Brazilian Institute of Geography and Statistics estimative to Brazilian population in the year 2019 [17].

## Discussion

Brazil is the fifth largest country in the world, with an area of 8,514,876 square kilometres divided into 27 federative units (26 states and 1 federal district), and a total of 5,570 cities [17]. Partly because of its large dimension, Brazil is a very heterogeneous country, with wide regional differences related to climate, lifestyle habits, socioeconomic characteristics, human development, social vulnerability, rates of inadequate water supply and sewage, population density across the states, and education level of the population [25]. Therefore, a single curve in the number of cases over time would not be representative of the whole country, due to a slower spread to the interior in some federative units. Thus, our main results are presented according to the five regions that comprise it.

Despite being the last of the five Brazilian regions to register a first case of COVID-19, the North region was one of the first to have a city with a health system overload. Currently, this is the case of Manaus, the capital of AM, where the mayor acknowledged a “collapse in the health system” and many patients are not getting access to proper health care, several of them dying at home [26]. This situation is even worse because Manaus is the only city in AM (the biggest state in the country) that has beds in intensive care units and, therefore, concentrates all intensive care in the state. This state also has few roads, and transport is mainly by rivers. The high incidences of COVID-19 in AM and AP, seen on 15 May, place these two states at the top of national ranking, as compared to states with the highest incidence rates in other regions. In addition, four of the seven states in the North, including AM and AP, present high SVI scores, and the worst rate of hospital beds per population over the country is found precisely in these two states, which would partly explain their worse performance, a chaotic situation in health assistance in Manaus and why the North has shown the highest incidence of COVID-19 among Brazilian regions until 15 May 2020.

The fight against COVID-19 becomes even more worrying because AM has a population of approximately 168,000 indigenous natives, representing 20% of the total indigenous population in Brazil. These populations are known to be more susceptive to epidemics and vulnerable to the occurrence of health conditions in general due to unfavourable socioeconomic conditions [27]. Furthermore, indigenous communities usually live in isolated areas, far from medical posts, doctors, and access to basic medicines [26].

Northeast is the second most populous region in the country. Although social isolation rates in the Northeast states on 15 May were among the highest of the entire country, the region also has the second highest number of cases and deaths by COVID-19. Two states in this region, CE and PE, had incidence increases and lethality rates due to COVID-19 in 15 May higher than SP, the state from Southeast region with the highest numbers cases and deaths due to this disease. It is important to note that both CE and PE have higher SVI scores and inadequate water supply and sewerage rates than SP, which may explain, at least in part, those differences. These observations suggest that socioeconomic context and social inequality play a role in the spread of COVID-19 in Brazil, as observed in the North. Supporting this hypothesis, states in the North and Northeast with higher SVI scores showed greater increases in the incidence rate from 14 April to 15 May. Khalatbari-Soltani et al. (2020) highlight the role that socioeconomic characteristics may play in transmission, incidence and health outcomes in COVID-19, as well as the need for high-quality data on socioeconomic factors, which can be useful in the development and implementation of public health actions [28].

The Southeast region includes the three largest metropolitan areas in Brazil, that is, the cities of São Paulo, Rio de Janeiro and Belo Horizonte; and the main industrialized areas. It is also the region with the biggest population among the five Brazilian political regions, as well as the one with the highest numbers of cases and deaths by COVID-19. The first case reported in Brazil, in São Paulo city, occurred on a date very close to the Carnival party (21 to 26 February 2020). The arrival and the beginning of the spread of COVID-19 in the Brazilian capitals may have occurred at the time of this party.

Brazilian carnival is well known all over the world and attracts many foreigners to the main cities of the country. In this traditional Brazilian party, most of the population takes to the streets in immense agglomerations, to the sound of typical local rhythms, mainly samba. Although Carnival took place before the first reported case of COVID-19 in Brazil, many people may have been infected at that time and some of them may have had asymptomatic illness or mild flu, which was not reported.

South region has the lowest numbers of cases and deaths, as well as the lowest incidence rates of COVID-19, along with the Midwest region, which may be reflect the fact that both regions have the smallest populations among the five Brazilian political regions. At least in the South, this may be explained, in part, by the rate of inadequate water supply and sewerage, less than 1% in all three states, and by the low scores on SVI. There are concerns about the effects of winter, which is more pronounced in this region and whose coldest months are just ahead (May to August). While data on SARS-CoV-2 seasonality is not yet clear, routine diagnostics records show a strong and consistent seasonal variation of the four previous coronavirus epidemics (229E, HKU1, NL63, OC43) [29]. In this line, a study based on data from 130 Chinese cities, between 20 January and 2 March 2020, pointed out that lower temperature and humidity are likely to favour the transmission of COVID-19 [30]. Thus, concerns about increased spread during the winter months seem to make a lot of sense. On the other hand, a recent study evaluating meteorological factors related to the spread of COVID-19, conducted in five cities of Brazil, showed it is favoured by higher mean temperatures (27.5°C) as well as intermediate relative humidity (in the range of 80%), although the study was carried out entirely before winter in Brazil [31]. Although these meteorological conditions are compatible with those observed in Manaus, AM, in the North, in this study we demonstrated that other factors, such as health indicators, are more important to explain the greater numbers of cases and deaths in this region.

Two states in the Midwest, MT and MS, had reported less than 800 cases of COVID-19 on 15 May 2020, while GO had 1,572 cases on the same date. These three states had low incidence rates, ranging 17.24 to 22.40 per 100,000 individuals. DF is an exception in Midwest region, with the highest number of cases (3,787) and incidence rate (125.59 per 100,000). The lower numbers of cases in MT and MS may be related to the presence of a large rural area and to the closure of borders between federative units, since these states presented the lowest social isolation rates in the county (less than 40%). On the other hand, as it contains the Brazilian capital (Brasília), DF has a greater circulation of people compared to the rest of the Midwest region, with a greater number of both national and international flights, which may explain part of this higher incidence within Midwest.

We believe that, in Brazil, cases of COVID-19 may be underreported, since the number of diagnostic tests available is much lower than necessary. In addition, testing for COVID-19 has been reduced only to people admitted to hospitals, and yet it is not enough for everyone. Many Brazilians died in their homes, which may cause fatal cases of COVID-19 to be underreported in our country. No widespread testing on all suspected cases has been performed. In a population-based study conducted in the state of RS, in April 2020, reported cases represented only around 10% of all detected cases [32]. In addition, states registries show higher numbers of deaths than Ministry of Health reports, which may indicate that fatality cases are underestimated. According to this data, since the epidemic beginning in Brazil, registries reported 58,839 deaths caused by SARS, respiratory failure and pneumonia, and only 12,344 deaths by COVID-19. In this regard, other studies have highlighted the potential for undocumented cases and low estimation due to limited testing in China to the spread of SARS-CoV-2 [33], and also in Brazil whose numbers of cases and deaths can be much higher than the official data released [34].

The main results of our study are in line with other previously published reports, showing that disadvantaged socioeconomic regions and poor health indicators perform worse during sanitary crises, with higher risks of disease and mortality [28, 35]. There is no reason to believe that the COVID-19 pandemic is an exception; as we showed here, regions of poor health determinants performed worse against the disease.

In low-income populations, where absolute poverty is a major problem, access to basic needs, such as clean water, sanitation and food, will have a substantial impact on how easily people can practice any social distancing measures. Furthermore, in low- and middle-income regions, people are more likely to live in overcrowded homes or neighbourhoods, making it impossible to isolate elderly or vulnerable people [28].

Before COVID-19 pandemic, Brazil was already facing an economic crisis, with 11.6 million unemployed in the fourth quarter of 2019 [36]. As of March 2020, during the outbreak of the SARS-CoV-2, there were more than 38 million informally self-employed people in Brazil, with no Social Security coverage or any other forms of protection [37]. Although emergency financial support has been settled by Federal Government for informal workers, the amount granted is less than 60% (about $ 110 US dollars) of the minimal income established in the country. This circumstance may have reduced the rate of social isolation, especially among the lower middle-and low-income people. A delay in the implementation of social policies leads to an aggravation of this health crisis and we believe it may also have influenced people to a lower adhesion to social isolation and, consequently, to the increase in the spread of COVID-19.

Furthermore, the Brazilian Federal Government took a stand against WHO recommendations regarding social isolation during COVID-19 pandemic, with several speeches neglecting the severity of the disease and the need for social isolation, in favour of the country’s economic development. We believe that these speeches may also have influenced people to a lower adhesion to social isolation and, consequently, to the increase in cases and deaths due to COVID-19.

When the history of COVID-19 is told, Brazil will likely be remembered as the country with the highest number of cases in Latin America (330,890 cases with 21,048 deaths as of 22 May) [16]. Unfortunately, this sad historical data will describe countless deaths and a drastic increase in unemployment and hunger, as well as each measure taken by the governments of the countries affected by COVID-19. During this pandemic, people in Brazil find themselves divided between two different guidelines on what they should do: on the one hand, a strong campaign by the media, the minister of Health, governors and mayors, urging people to maintain social isolation and stay home. On the other hand, the attitudes of the President of Brazil, sowing confusion, openly disrespecting and discouraging sensible measures of physical distancing and lockdown, and stimulating frequent political crises during this period, as very well described and documented in a recent editorial published in The Lancet [38].

The limitations of this study include those related to incomplete information from government databases, which depend on inputs from thousands of different cities about cases and deaths, and the data of Brazilian last census, carried out in 2010. In addition, there is a restriction of tests for SARS-CoV-2 in the Brazilian population, which produces underestimated information.

Data presented in 15 May 2020 demonstrate that Brazil is still far from reaching the epidemic peak and suggest that actions to support the disadvantaged population by the government should be improved, since a large number of Brazilians live in poverty, in small houses with a great number of people, who share the same rooms, where social isolation is unfeasible. Despite this, in line with WHO recommendations, we believe that social isolation should be encouraged by the government and employers as far as possible, in order to prevent the spread of COVID-19 and further increases in cases and deaths.

## Data Availability

All data are fully available without restriction.
It is SPSS database.

## Acknowledgements

EMG-S thanks the Universidade Federal Mato Grosso do Sul (UFMS) for the opportunity of being a visiting professor (contract PROGEP/RTR No 2019/000230). This study was partly financed by the Coordination for the Improvement of Higher Education Personnel - Brazil (Coordenação de Aperfeiçoamento de Pessoal de Nível Superior (CAPES)), through the granting of scholarships to postgraduate students. We thank the Federal University of Mato Grosso do Sul (UFMS) for the support for language revision and payment of publication charge by EDITAL PROPP / UFMS N° 143/2019.

## Authors’ contributions

EMG-S designed the research study, analysed the data, performed statistical analysis, and wrote the paper. CP, GWG, SSW, EJS and TBRG collaborated in the writing of the manuscript. Acquisition of data and references, EJS, ICP, KGJ, RFAZ, and AGB. Analysis and interpretation of data were also performed by EMG-S, SSW, EJS, CP, and GWG. Critical revision, EMG-S, CP, GWG, TBRG, SSW, and ADC.

---

**S1 Figure.**
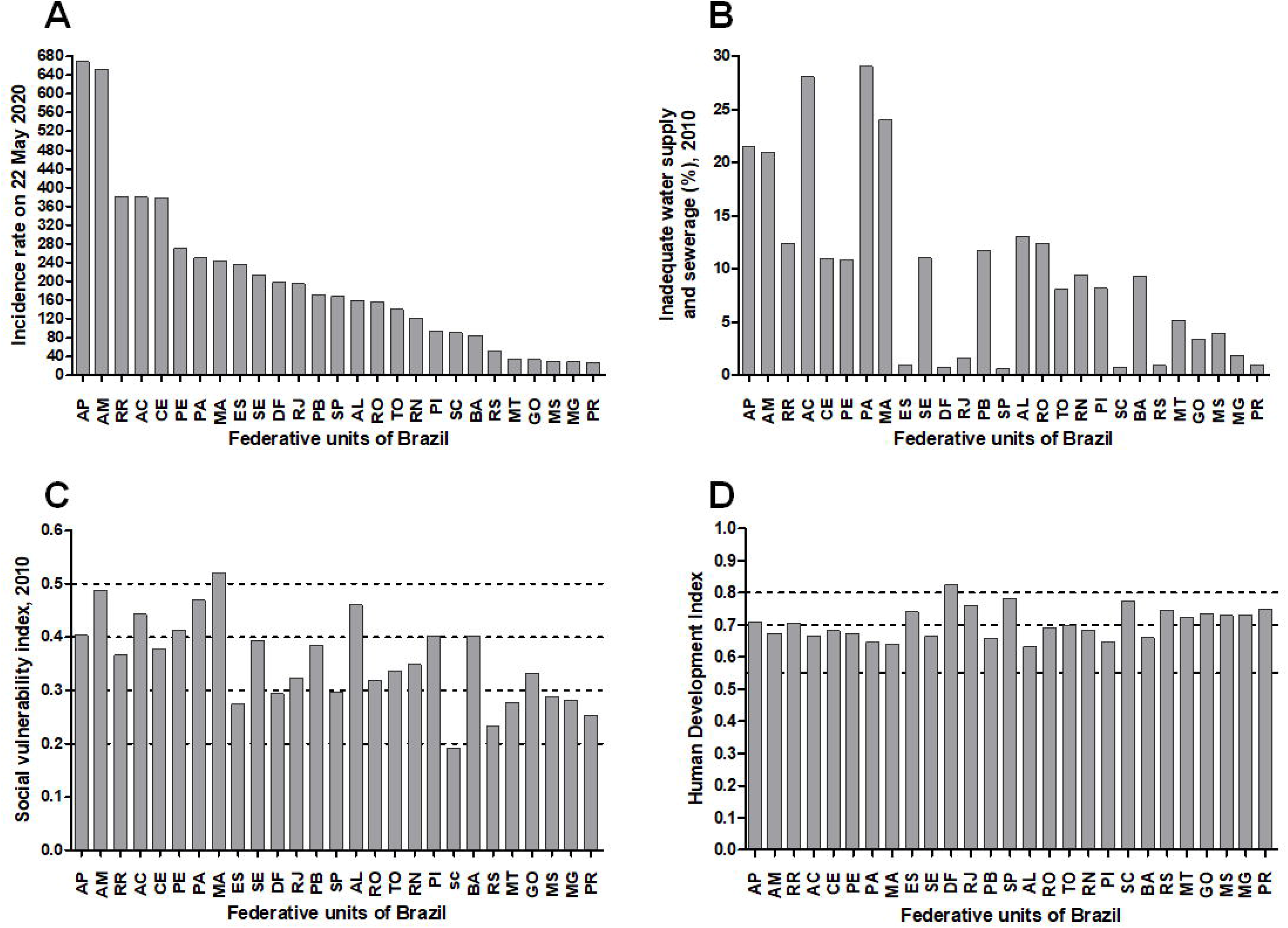
**(A) Incidence rate of COVID-19 on 22 May 2020, (B) Percentage of inadequate water supply and sewerage, (C) Social Vulnerability Index (SVI), and (D) Human Development Index (HDI) in the Brazilian federative units. Figure B, C and D show federative units in decreasing order of COVID-19 incidence, according to data from 22 May 2020 (A). Lines in Figure 1C represents values of SVI classified as very low (0 to 0.200), low (0.201 to 0.300), medium (0.301 to 0.400), high (0.401 to 0.500), and very high (0.501 to 1) [22]. Lines in Figure 1D represents values of HDI classified as very high (≥ 0.800), high (0.700 to 0.799), medium (0.550 to 0.699) and low (< 0.550) [21]**. AC, Acre; AL, Alagoas; AP, Amapá; AM, Amazonas; BA, Bahia; CE, Ceará; DF, Distrito Federal; ES, Espírito Santos; GO, Goiás; MA, Maranhão; MT, Mato Grosso; MS, Mato Grosso do Sul; MG, Minas Gerais; PA, Pará; PB, Paraíba; PR, Paraná; PE, Pernambuco; PI, Piauí; RJ, Rio de Janeiro; RN, Rio Grande do Norte; RS, Rio Grande do Sul; RO, Rondônia; RR, Roraima; SC, Santa Catarina; SP, São Paulo; SE, Sergipe; TO, Tocantins.

**S2 Figure.**
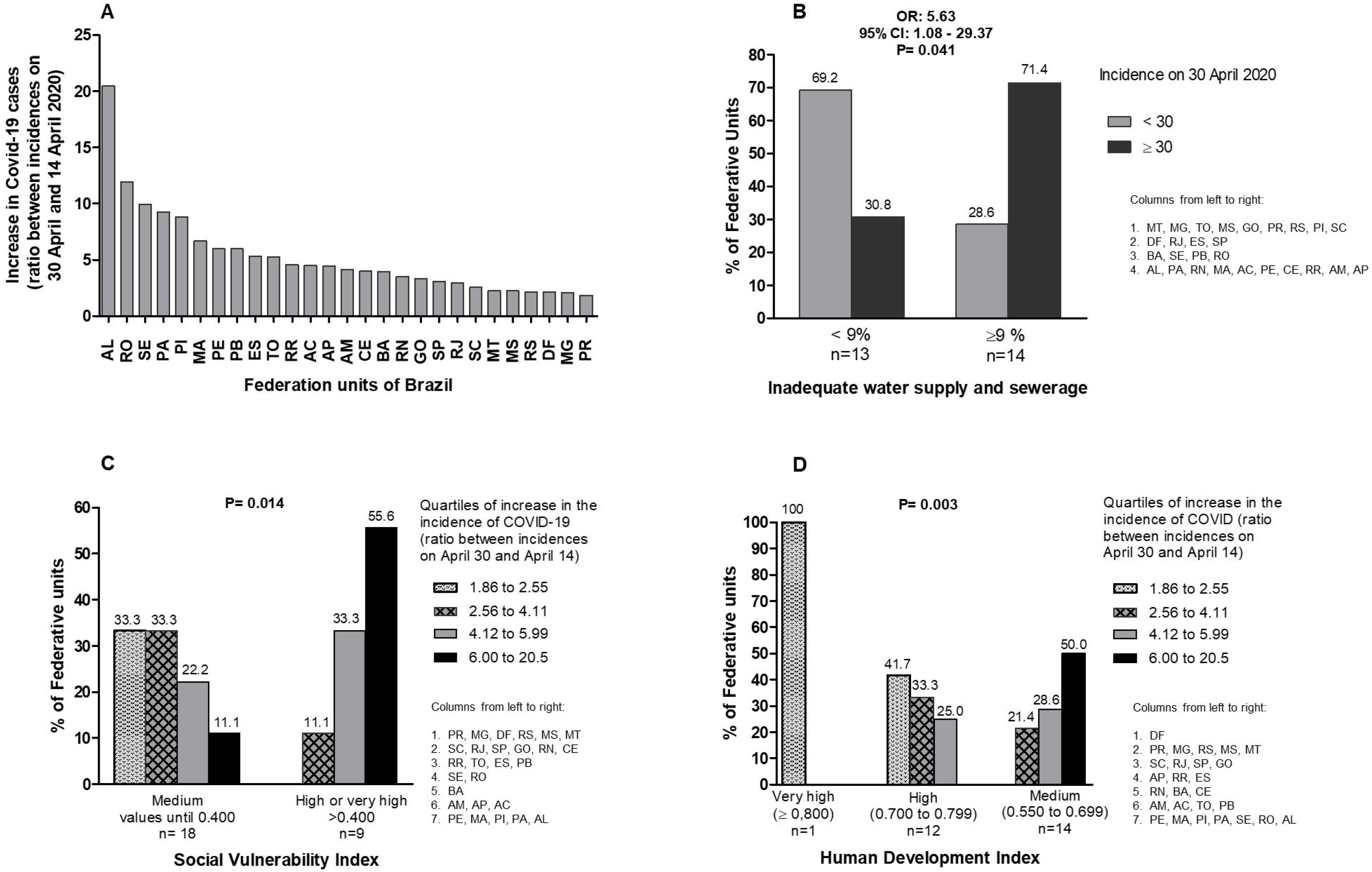
**Increase in cases over a period of 16 days in April, using the ratio between incidences per 100,000 on 30 and 14 April 2020. B. Chi-square test. C and D. Likelihood ratio**. AC: Acre, AL: Alagoas, AP: Amapá, AM: Amazonas, BA: Bahia, CE: Ceará, DF: Distrito Federal, ES: Espírito Santos, GO: Goiás, MA: Maranhão, MT: Mato Grosso, MS: Mato Grosso do Sul, MG: Minas Gerais, PA: Pará, PB: Paraíba, PR: Paraná, PE: Pernambuco, PI: Piauí, RJ: Rio de Janeiro, RN: Rio Grande do Norte, RS: Rio Grande do Sul, RO: Rondônia, RR: Roraima, SC: Santa Catarina, SP: São Paulo, SE: Sergipe, TO: Tocantins.

